# Web and social media searches highlight menstrual irregularities as a global concern in COVID-19 vaccinations

**DOI:** 10.1101/2022.01.30.22270125

**Authors:** Ariel Katz, Yoav Tepper, Ohad Birk, Alal Eran

## Abstract

**Background:** Delineation of public concerns that prevent vaccine compliance is a major step in generating assurances and enhancing the success of COVID-19 prevention programs. We therefore sought to identify public concerns associated with COVID-19 vaccines, as reflected by web and social media searches, with a focus on menstrual irregularities.

**Methods:** We used trajectory analyses of web and social media search data in combination with global COVID-19 data to reveal time-dependent correlations between vaccination rates and the relative volume of vaccine and period related searches.

**Results:** A surge of *period and vaccine* related Google searches followed the introduction of Covid vaccines around the world, and the commencement of vaccination programs in English speaking countries and across the United States. The relative volume of searches such as “Covid vaccine menstrual irregularities”, “Covid vaccine menstrual period”, “Pfizer vaccine menstruation”, and “Moderna vaccine menstruation” was each significantly correlated with vaccination rates (Spearman r = 0.42-0.88, P = 4.33 × 10^−34^-1.55 × 10^−5^), and significantly different before and after the introduction of COVID vaccines (Mann-Whitney P = 2.00 × 10^−21^-7.10 × 10^−20^). TikTok users were more engaged in period problems in 2021 than ever before.

**Conclusions:** International, national, and state-level correlations between COVID-19 vaccinations and online activity demonstrate a global major concern of vaccine-related menstrual irregularities. Whether it is a potential side effect or an unfounded worry, monitoring of web and social media activity could reveal the public perception of COVID-19 prevention efforts, which could then be directly addressed and translated into insightful public health strategies.

## Introduction

Women around the world have reported a link between COVID-19 vaccinations and changes in their menstrual cycle regularity and intensity^1^. One of the early reports was a tweet by Kathryn Clancy, a Professor of Anthropology at the University of Illinois, which was followed by hundreds of responses of women identifying with her concerns, and in many cases reflecting on having similar menstrual irregularities following vaccination. Later reports on menstrual irregularities following vaccination were met with criticism^2^, some suggesting that pandemic-related stress might be the cause of irregularities. Antivaxxers later highlighted the worries regarding possible vaccine-induced menstrual irregularities, expanding unverified and often manipulative concerns, and promoting fears of vaccine– associated abortions and infertility^3,4^. Recent studies have examined the effects of COVID-19 disease^5^ and vaccines^6–8^ on menstrual irregularities, suggesting lack of adequate reporting of such irregularities due to women’s reluctance to discuss these matters with their physicians^9^. Yet the public perception of a potential link between COVID-19 vaccines and menstrual irregularities remains largely unknown. To examine the spread and magnitude of this concern we mined web and social media search data and its relation to the specific timing of national and state-level COVID-19 vaccination drives.

In doing so, we utilized Google Trends^10^, a powerful tool for analyzing population behavior and search trends^11^. Google Trends is capable of summarizing time and location-specific searches, enabling integrative spatiotemporal analyses, including those related to health^12^. For example, recent infodemiological studies have leveraged Google Trends to model disease prevalence^13^ and symptomatology^14^, especially in the context of COVID-19^15-17^.

## Methods

We used Google Trends to examine the relative volume of searches for “Covid vaccine menstrual irregularities”, “Covid vaccine menstruation”, “Covid vaccine menstrual period”, “vaccine and period”, “Pfizer vaccine menstruation”, and “Moderna vaccine menstruation” worldwide, in English speaking countries and across the US between January 2020 and November 2021. Specifically, we used the “search term” functionality of Google Trends with no category. The settings of all analyses are recorded in all results files available at https://bit.ly/covid_vaccine_period_search_data.

We also mined TikTok, a social network serving more than a billion users, 53% of whom are female and 78% of whom are under the age of 2418. Using Analisa.io (https://analisa.io), we examined the activity of “#periodproblems” from January 2019 to October 2021.

We integrated the search data with COVID-19 vaccination data obtained from Our World In Data (https://ourworldindata.org/covid-vaccinations) and other COVID-19 aggregated statistics, including the number of cases and deaths, obtained from https://github.com/CSSEGISandData/COVID-19/tree/master/csse_covid_19_data. Our python code that performs and visualizes all analyses is freely available at https://github.com/SgtTepper/CovidTrends. Specifically, it mines Google Trends via pytrends, integrates the results with COVID statistics using pandas, computes Spearman correlations and Mann Whitney U tests using SciPy stats, plots co-occurring multimodal data on the same timeline using seaborn, and plots relative search volume by state using plotly.

Our statistical analyses focused on assessing the relations between The Google Trends search volume index (SVI) and the normalized vaccination rate (NVR) for each search term of interest. SVI measures the relative popularity of a certain search at specific locations and during specific times. This relative index is normalized from 0 to 100, all with respect to the prespecified time interval and locations. NVR reflects the daily total of vaccinated individuals in the studied location, normalized such that the maximal number in the examined time interval is 100. In other words, it is 100*daily total of vaccinated individuals / maximal daily total of vaccinated individuals in the studied period. We utilized two complementary analytical approaches. First, we used Spearman correlation between SVI and NVR across the entire studied period to assess the monotonic relationships between SVIs of interest and the NVR for Covid in the studied location. Second, we used Mann-Whitney U tests to examine the significance of the difference in the distribution of SVI before and after the introduction of Covid vaccines in the studied location.

## Results

Around the world, the relative volume of Google searches for “Covid vaccine menstrual irregularities”, “Covid vaccine menstrual period”, “vaccine and period”, “Pfizer vaccine menstruation”, “Moderna vaccine menstruation”, and similar phrases grew dramatically following the initiation of COVID-19 vaccination drives (**Figure 1**). There was a significant correlation between the global NVR and the worldwide relative SVI of “Covid vaccine menstrual irregularities” (Spearman r = 0.86, P = 2.70 × 10^−20^), and a significant difference in SVI distributions of this term before and after the commencement of vaccination programs (Mann-Whitney U P = 7.10 × 10^−20^). Top related queries show that Google users were mostly concerned with menstrual irregularities after a Covid vaccine (**Figure 1a**). Similarly, we found a significant correlation with the global NVR and a significant difference in SVI distributions before and after the introduction of vaccines for the term “vaccine and period” (Spearman r = 0.86, Spearman P = 2.01 × 10^−30^, Mann-Whitney U P = 7.10 × 10^−20,^ **Figure 1b**). We specifically used such non-medical term to reflect common language used by young females, which represent much of our target population, namely women of reproductive age authorized to receive a COVID vaccine (i.e. between 16 and 50 years of age). The top related queries show that users are referring to covid vaccines and period after such vaccine (**Figure 1b**). We also examined the relative search dynamics of “Covid vaccine menstrual period” as a more specific term (**Figure 1c**). Its SVI was similarly correlated with the global NVR (Spearman r = 0.86, Spearman P = 2.78 × 10^−30^) and significantly different before and after the introduction of vaccines (Mann-Whitney U P = 7.10 × 10^−20^)

**Figure 1:**
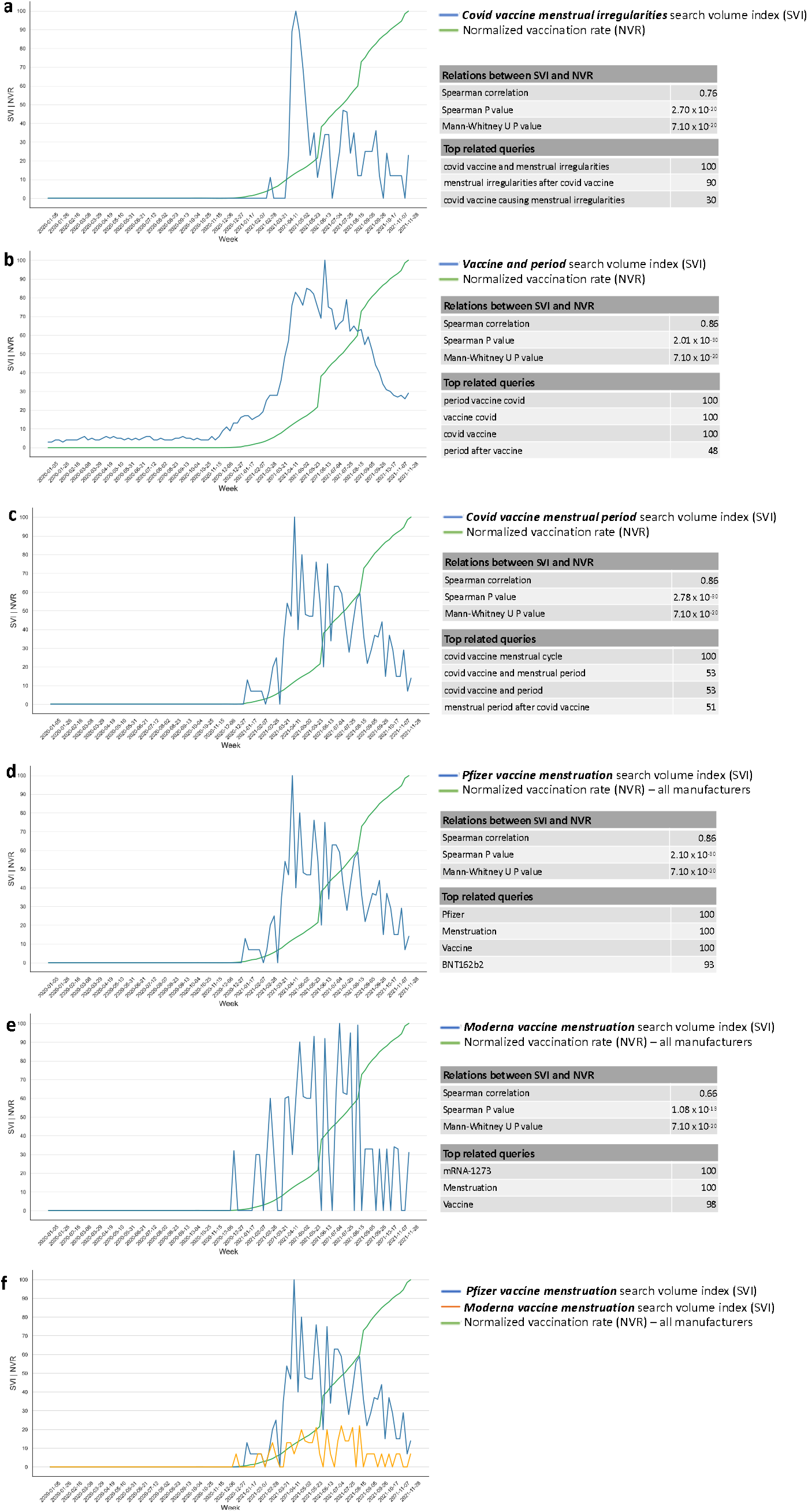
Relative volume of Google searches and their temporal relation to global vaccination rates for the terms (**a**) “Covid vaccine menstrual irregularities”, (**b**) “Vaccine and period”, (**c**) “Covid vaccine menstrual period”, (**d**) “Pfizer vaccine menstruation”, and (**e**) “Moderna vaccine menstruation”. (**f**) A direct comparison of “Pfizer vaccine menstruation” and “Moderna vaccine menstruation” search behavior and their relations to the global NVR. For each search term, the Spearman correlation between the search volume index (SVI) and normalized vaccination rate (NVR) is shown, as well as the significance of the difference in SVI distributions before and after the introduction of vaccines. Top related queries are also shown for context. The full lists of related queries and entities for each search term is available at https://bit.ly/covid_vaccine_period_search_data

Next, focusing on leading COVID vaccine manufacturers, we found a significant correlation with the NVR and a significant difference in SVI distributions before and after the commencement of vaccination drives for the searches “Pfizer vaccine menstruation” (Spearman r = 0.86, Spearman P = 2.10 × 10^−30^, Mann-Whitney U P = 7.10 × 10^−20,^ **Figure 1d**) and “Moderna vaccine menstruation” (Spearman r = 0.66, Spearman P = 1.08 × 10^−13^, Mann-Whitney U P = 7.10 × 10^−20^, **Figure 1e**). Of note, the vaccination data used was blind to the manufacturer and included all COVID vaccines recorded. A head-to-head comparison of the SVIs of “Pfizer vaccine menstruation” and “Moderna vaccine menstruation” showed that the Pfizer search term was more common than the Moderna one, consistent with the wider use of Pfizer’s BNT162b2 as compared to Moderna’s mRNA-1273 around the world (**Figure 1f**).

To ensure that no one country accounts for the observed global relations in search behavior and vaccination rates, we next repeated the same analyses in four English speaking countries. For example, the relative volume of Google searches for the combination of terms “period” and “vaccine” peaked following the initiation of COVID-19 vaccination drives, with a later 3-5-fold further increase once ∼50% of the population has been vaccinated (**Figure 2**). Country-specific timing of surges in “vaccine and period” related searches and its temporal relation to the local vaccination drive control for possible effects of geographic location and the local culture. Note that the analyses focused on countries sharing English as the main language, ensuring the searches were done mostly with similar wording and adequate spelling. Figure 2 also shows the top related queries in each country, demonstrating that users were mainly concerned with period after Covid vaccine.

**Figure 2:**
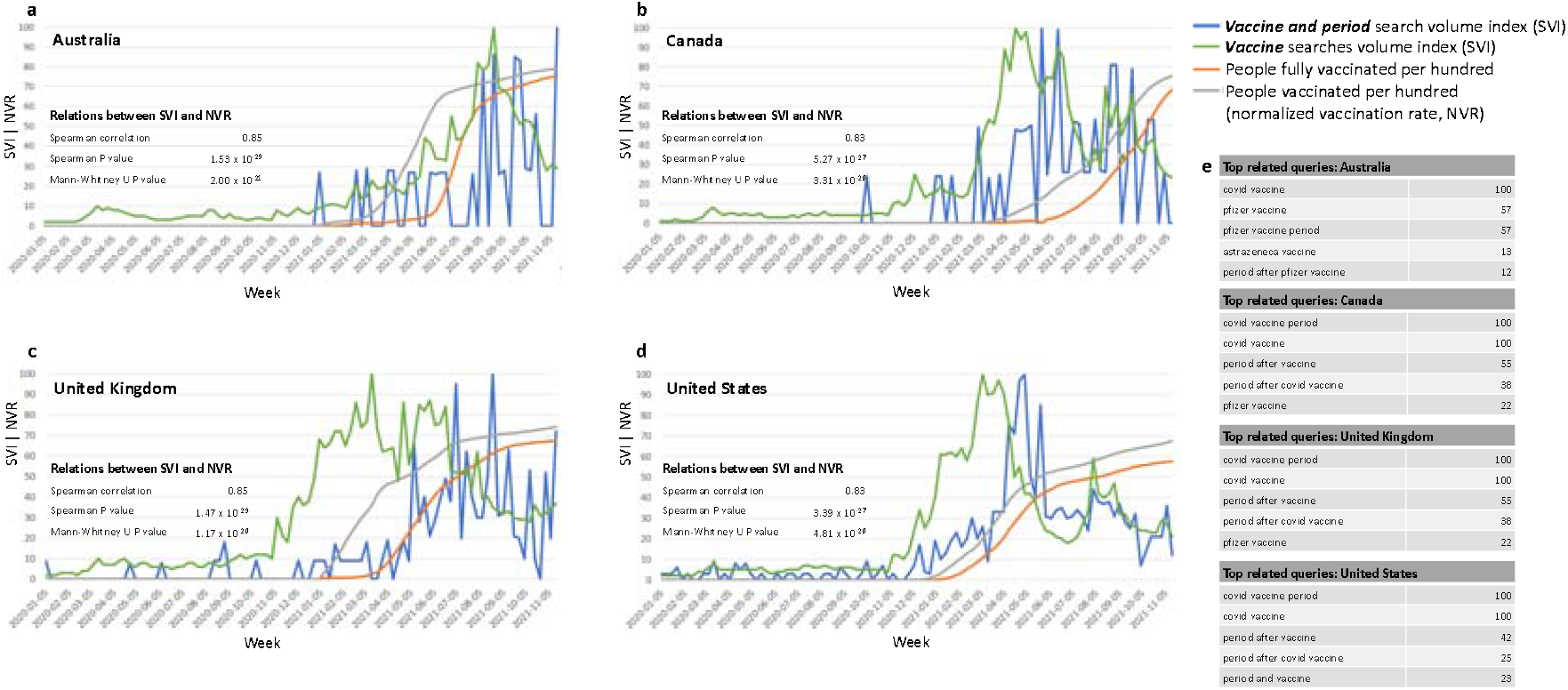
Relative volume of Google searches for the combination of terms “period” and “vaccine”, and their temporal relation to national vaccination rates in English speaking countries. (**a**) Australia, (**b**) Canada, (**c**) United Kingdom, (**d**) United States. The volume of “vaccine” searches is shown as a positive control. The Spearman correlation between the relative search volume and national normalized vaccination rates is shown for each country, as is the significance of the difference in SVI distributions before and after the country-specific commencement of vaccination drives. (**e**) The top related queries in each country demonstrate the relevance of this non-medical, common search term. The full lists of related queries and entities for each search term is available at https://bit.ly/covid_vaccine_period_search_data.

Another demonstration of the ubiquitous change in search behavior and its temporal relation to country-specific vaccination drives is exemplified by the dynamics of the more specific term “Covid vaccine menstruation” in four English-speaking countries (**Figure 3**). Its related queries demonstrate that users were mostly concerned with the impact of Covid vaccines on menstruation (**Figure 3e**).

**Figure 3:**
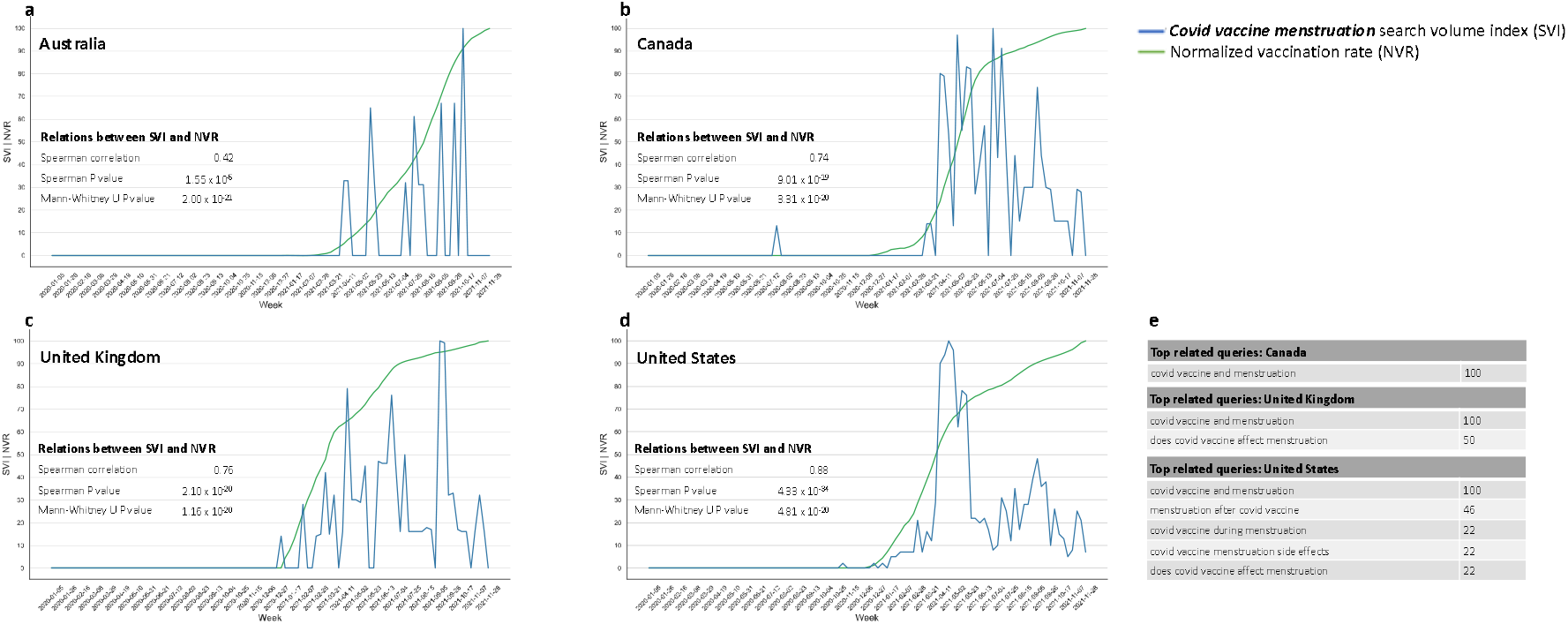
Search behavior for “Covid vaccine menstruation” and its temporal relation to national vaccination rates in English speaking countries. (**a**) Australia, (**b**) Canada, (**c**) United Kingdom, (**d**) United States. The Spearman correlation between the relative search volume (SVI) and national normalized vaccination rates (NVR) is shown for each country, as is the significance of the difference in SVI distributions before and after the country-specific commencement of vaccination drives. (**e**) The top related queries in each country demonstrate the search context of this term. Note that no related queries were found in Australia. The full lists of related queries and entities for each search term is available at https://bit.ly/covid_vaccine_period_search_data.

Similar relations were detected within the United States (**Figure 4**). Overall, Google searches for the combination of “period” and “vaccine” were frequent between October 2020 and October 2021, to a somewhat lesser extent in the mid-U.S (**Figure 4a**). As exemplified in Figure 4b-4m, searches for the combined terms “period” and “vaccine” became extremely prominent with the initiation of COVID-19 vaccines in each state. Notably, further major peaks in searches for the terms “period” and “vaccine” appeared 2-6 months following initiation of vaccines, suggesting that searches did not only reflect worries, but were possibly triggered by menstrual irregularities following vaccines.

**Figure 4.**
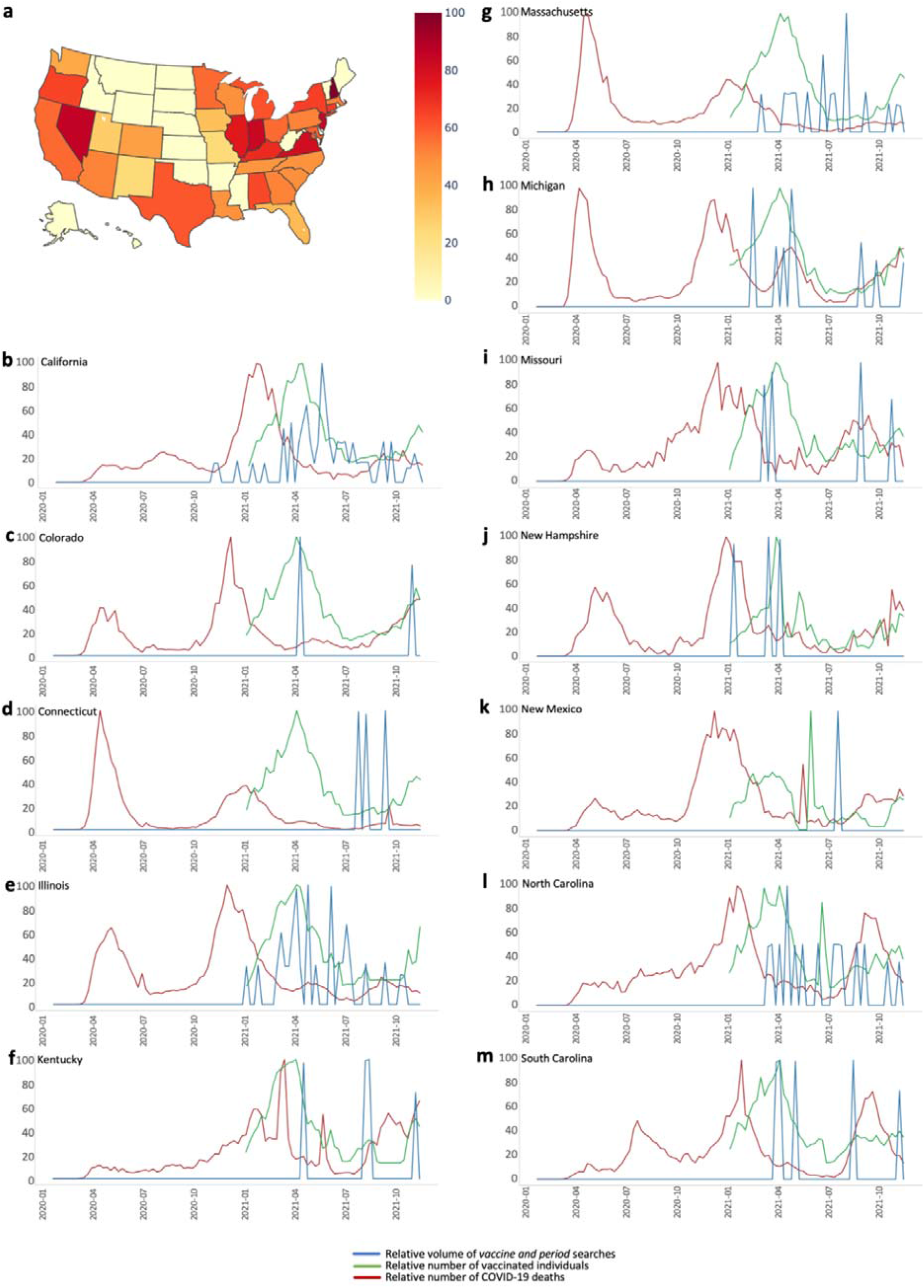
State-level “vaccine and period” searches and their temporal relation to vaccination rates. (**a**) Relative volume of “vaccine and period” related searches across the United States between October 1^st^ 2020 and October 1^st^ 2021. (**b-m**) Temporal relations of “vaccine and period” searches and state-specific vaccinations and deaths, normalized to their maxima within the studied period, namely 10/1/2020 – 10/1/2021 in (**b**) California, (**c**) Colorado, (**d**) Connecticut, (**e**) Illinois, (**f**) Kentucky, (**g**) Massachusetts, (**h**) Michigan, (**i**) Missouri, (**j**) New Hampshire, (**k**) New Mexico, (**l**) North Carolina, (**m**) South Carolina. Similar timelines for all 50 states are available at https://bit.ly/covid_vaccine_period_search_data. Of note, COVID-19 vaccinations started in December 2020 in California, while state-level vaccination statistics are available from January 2021. A possible explanation for the minor increase in search volume in November 2020 in California is the Phase III clinical trial of COVID-19 vaccine by both Pfizer and Moderna, which included thousands of California residents^19-21^.

Moreover, analysis of worldwide use and reactions to “#periodproblems” in the video-focused social network TikTok between January 2019 and October 2021 (**Figure 5**) demonstrated an initial significant peak of this hashtag in March 2020 (both in the number of exposures and the number of responses to the term), possibly reflecting pandemic stress. Yet this peak subsided and dramatically re-emerged in January 2021with several major peaks, coinciding with the timing of initiation of vaccination programs around the world.

**Figure 5.**
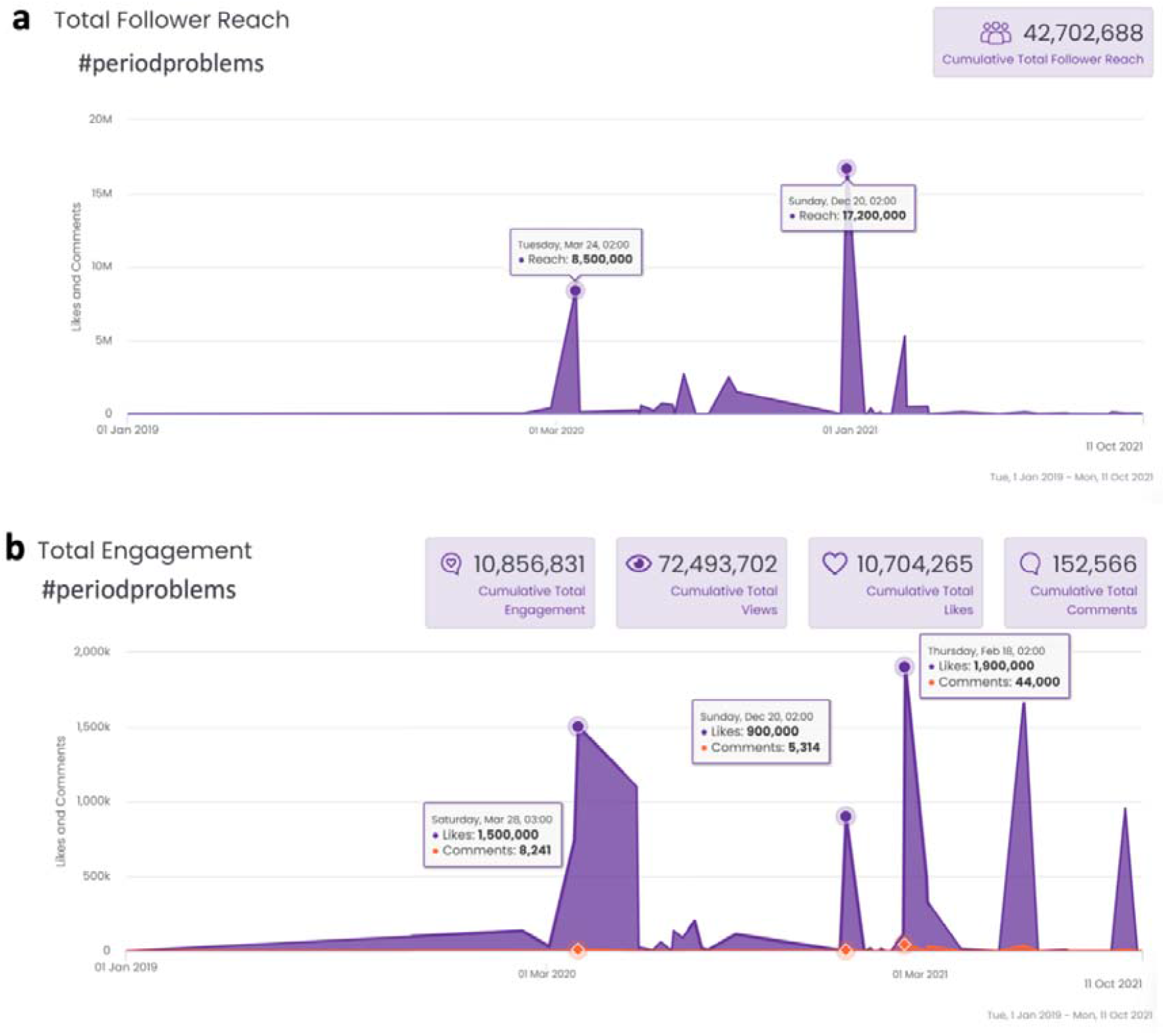
Worldwide TikTok use of the hashtag #periodproblems between January 2019 and October 2021. There is an initial dramatic peak of this hashtag (both in number of exposures to this term and number of responses to the term) in March 2020, possibly reflecting pandemic stress, yet this peak subsided and dramatically re-emerged as of January 2021with several peaks, possibly reflecting timing of initiation of vaccines in different countries. Note the massive engagement in this topic during worldwide vaccination drives. (**a**) Exposures to TikTok videos containing “#periodproblems”. (**b)** Responses to TikTok videos containing “#periodproblems”.

## Discussion

This study finds a surge of Google searches related to COVID vaccine and the menstrual cycle following the initiation of national and state-level COVID-19 vaccination drives. While it may suggest that women of reproductive age vaccinated for COVID-19 might have experienced menstrual irregularities, it could also reflect the mere concerns of those not yet vaccinated. In either case this study does not support any link between COVID-19 vaccines and menstrual irregularities; Rather it documents the evolving public perception of such potential link. We also find that TikTok hashtag #periodproblems peaked at the beginning of the pandemic in March 2020, and again with the initiation of vaccination programs, early in 2021. This reflects menstrual irregularities being a significant worry associated with COVID-19 vaccinations, yet practically rules out claims that concerns about the menstrual cycle are merely an effect of pandemic-related stress.

One important limitation of this study is that it measures changes in relative search volumes, and not absolute values of users showing concerns. Third-party verification methods such as questionnaires and health care providers’-initiated surveys could be used to obtain a better understanding of the public perception of relations between COVID vaccination and menstrual irregularities.

The safety of COVID-19 vaccines in pregnancy and lack of discernable effects on fertility have been recently shown in several reports^22,23^. Studies of the possible effects of COVID-19 vaccines on menstruation have been initiated through questionnaires, doctors’ reports and other experimental strategies^24^. However, these studies encompass limited amounts of data in comparison to that available by monitoring web and social media activity. Moreover, studies based on physicians’ reports are lacking since women do not often report menstrual irregularities to their physicians, because of discomfort or inconvenience of going to the doctor, especially during the pandemic^9^. Our findings reflect menstrual cycle irregularities as a major worry of women in considering compliance with vaccination programs, suggesting possible effects of the vaccine on such irregularities, and highlight a clear need for scientifically sound studies testing possible effects of the vaccines on menstruation. Of note, our analysis deals with relative search volumes. Thus, significant changes in other searches at the same time and place might mask or distort the results.

This study highlights the use of an array of social media big data analyses, combining Google Trends and TikTok (representing a significantly younger cohort), in identifying public trends preventing vaccine compliance across different states and countries. Prior studies proved retrospectively through Google trends facts that were previously known, such as the initiation of the pandemic or effects of COVID-19 disease on smell. In contrast, this study highlights social media big data analysis as an effective massive-scale tool in the initial identification and determination of adverse effects (as well as positive effects) of the COVID-19 vaccine. Moreover, it verifies the findings across various states in the U.S. and other countries, ruling out effects of local trends, climates, publicities and regulations, as well as enables delineation of different effects of the various COVID-19 vaccines given in different states or countries. Thus, web and social media analyses enable improved strategies in identifying public health concerns and determining public health policies to improve vaccine compliance.

## Data Availability

All data produced are available online at https://github.com/SgtTepper/CovidSearchTrends/

## Data availability

All data and its visualizations generated as part of this study are available at https://bit.ly/covid_vaccine_period_search_data.

